# Assessing the Impact of the Covid-19 Pandemic on US Mortality: A County-Level Analysis

**DOI:** 10.1101/2020.08.31.20184036

**Authors:** Andrew C. Stokes, Dielle J. Lundberg, Irma T. Elo, Katherine Hempstead, Jacob Bor, Samuel H. Preston

## Abstract

**Background:** Covid-19 excess deaths refer to increases in mortality over what would normally have been expected in the absence of the Covid-19 pandemic. Several prior studies have calculated excess deaths in the United States but were limited to the national or state level, precluding an examination of area-level variation in excess mortality and excess deaths not assigned to Covid-19. In this study, we take advantage of county-level variation in Covid-19 mortality to estimate excess deaths associated with the pandemic and examine how the extent of excess mortality not assigned to Covid-19 varies across subsets of counties defined by sociodemographic and health characteristics.

**Methods and Findings:** In this ecological, cross-sectional study, we made use of provisional National Center for Health Statistics (NCHS) data on direct Covid-19 and all-cause mortality occurring in U.S. counties from January 1 to December 31, 2020 and reported before March 12, 2021. We used data with a ten week time lag between the final day that deaths occurred and the last day that deaths could be reported to improve the completeness of data. Our sample included 2,096 counties with 20 or more Covid-19 deaths. The total number of residents living in these counties was 319.1 million. On average, the counties were 18.7% Hispanic, 12.7% non-Hispanic Black and 59.6% non-Hispanic White. 15.9% of the population was older than 65 years. We first modeled the relationship between 2020 all-cause mortality and Covid-19 mortality across all counties and then produced fully stratified models to explore differences in this relationship among strata of sociodemographic and health factors. Overall, we found that for every 100 deaths assigned to Covid-19, 120 all-cause deaths occurred (95% CI, 116 to 124), implying that 17% (95% CI, 14% to 19%) of excess deaths were ascribed to causes of death other than Covid-19 itself. Our stratified models revealed that the percentage of excess deaths not assigned to Covid-19 was substantially higher among counties with lower median household incomes and less formal education, counties with poorer health and more diabetes, and counties in the South and West. Counties with more non-Hispanic Black residents, who were already at high risk of Covid-19 death based on direct counts, also reported higher percentages of excess deaths not assigned to Covid-19. Study limitations include the use of provisional data that may be incomplete and the lack of disaggregated data on county-level mortality by age, sex, race/ethnicity, and sociodemographic and health characteristics.

**Conclusions:** In this study, we found that direct Covid-19 death counts in the United States in 2020 substantially underestimated total excess mortality attributable to Covid-19. Racial and socioeconomic inequities in Covid-19 mortality also increased when excess deaths not assigned to Covid-19 were considered. Our results highlight the importance of considering health equity in the policy response to the pandemic.

**Author’s Summary:** *Why Was This Study Done?:*

- The Covid-19 pandemic has resulted in excess mortality that would not have occurred in the absence of the pandemic.
- Excess deaths include deaths assigned to Covid-19 in official statistics as well as deaths that are not assigned to Covid-19 but are attributable directly or indirectly to Covid-19.
- While prior studies have identified significant racial and socioeconomic inequities in directly assigned Covid-19 deaths, few studies have documented how excess mortality in 2020 has differed across sociodemographic or health factors in the United States.

*What Did the Researchers Do and Find?:*

- Leveraging data from 2,096 counties on Covid-19 and all-cause mortality, we assessed what percentage of excess deaths were not assigned to Covid-19 and examined variation in excess deaths by county characteristics.
- In these counties, we found that for every 100 deaths directly assigned to Covid-19 in official statistics, an additional 20 deaths occurred that were not counted as direct Covid-19 deaths.
- The proportion of excess deaths not counted as direct Covid-19 deaths was even higher in counties with lower average socioeconomic status, counties with more comorbidities, and counties in the South and West. Counties with more non-Hispanic Black residents who were already at high risk of Covid-19 death based on direct counts, also reported a higher proportion of excess deaths not assigned to Covid-19.

*What Do These Findings Mean?:*

- Direct Covid-19 death counts significantly underestimate excess mortality in 2020.
- Monitoring excess mortality will be critical to gain a full picture of socioeconomic and racial inequities in mortality attributable to the Covid-19 pandemic.
- To prevent inequities in mortality from growing even larger, health equity must be prioritized in the policy response to the Covid-19 pandemic.

## Introduction

The novel coronavirus disease 2019 (Covid-19) is an international public health emergency caused by the respiratory droplet transmission of the coronavirus-2 (SARS CoV-2) virus [1]. SARS CoV-2 infects humans through the lung epithelium and is associated with a high incidence of acute respiratory distress syndrome, vascular injury, and death [2]. The United States has emerged as an epicenter of the Covid-19 pandemic, with over 18.2 million confirmed cases and 322,218 deaths as of December 22, 2020 [3].

Vital registration data sourced from death certificates on cause of death are likely to underestimate the mortality burden associated with the Covid-19 pandemic for several reasons [4,5]. First, some direct deaths attributable to Covid-19 may be assigned to other causes of death due to an absence of widespread testing and low rates of diagnoses at the time of death [6]. Additionally, direct deaths from unfamiliar complications of Covid-19 such as coagulopathy, myocarditis, inflammatory processes, and arrhythmias may have caused confusion and led to attributions of death to other causes, especially early in the pandemic and among persons with comorbid conditions [7–9]. Second, Covid-19 death counts do not take into account the indirect consequences of the Covid-19 pandemic on mortality levels [10,11]. Indirect effects may include increases in mortality resulting from reductions in access to and use of health care services and psychosocial consequences of stay-at-home orders [12]. Increases in stress, depression, and substance use related to the pandemic could also lead to suicides and overdose deaths [13,14]. Economic hardship, housing insecurity, and food insecurity may cause indirect deaths, especially among those living with chronic illnesses or who face acute heath emergencies and cannot afford medicines or medical supplies [15–17]. On the other hand, the pandemic may reduce mortality as a result of reductions in travel and associated motor vehicle mortality, lower air pollution levels, or the possible benefits of Covid-19 mitigation efforts (i.e. mask wearing and physical distancing) on reducing influenza spread [18,19]. It is also possible that Covid-19 deaths are over-recorded in some instances, e.g., because some deaths that should have been assigned to influenza were instead assigned to Covid-19. Finally, the Covid-19 pandemic may reduce mortality from certain other causes of death because of frailty selection; those who die from Covid-19 may have been unusually frail and vulnerable to death from other diseases.Consequently, the rate of death from those diseases may decline and offset some of the increase in all-cause mortality attributable to Covid-19 deaths alone.

The term “excess deaths” refers to differences in mortality relative to what would have been expected in the absence of the Covid-19 pandemic. Typically, excess deaths are computed relative to a recent historical benchmark for the same population. Excess deaths include deaths directly assigned to Covid-19 on death certificates and excess deaths not assigned to Covid-19, which were either misclassified to other causes of deaths or were indirectly related to the Covid-19 pandemic. Using deaths from all causes to measure the excess mortality impact of the Covid-19 pandemic can help circumvent biases in vital statistics, such as low Covid-19 testing rates, reporting lags, and differences in death certification coding practices, and capture excess deaths indirectly related to the Covid-19 pandemic. As such, estimates of excess deaths from all causes associated with the pandemic provide a useful measure of the total mortality burden associated with Covid-19 [5].

Previous reports have estimated excess deaths in several different ways. The National Center for Health Statistics (NCHS) estimates excess mortality through comparison of mortality levels in 2020 to historical mortality data by week and geographic location [20]. They present a range of values for excess deaths based on different historical thresholds, including the average expected count or upper boundary of the uncertainty interval, and apply weights to the 2020 provisional death data to account for incomplete data. In contrast, Weinberger et al. and Woolf et al. use multivariable Poisson regression models to evaluate increases in the occurrence of deaths due to any cause across the US [21,22]. Weinberger et al. adjust for influenza activity [21]. Kontis et al. apply Bayesian ensemble modeling to obtain smoothed estimates of excess deaths by age and sex in the UK [23]. These studies make estimates for each state or country individually and do not allow a relationship between all-cause mortality and Covid-19 mortality to be identified through analysis across smaller area units such as counties. While prior research has documented significant racial and socioeconomic inequities in directly assigned Covid-19 deaths [24–29], few studies have documented how excess mortality in 2020 has differed across sociodemographic or health factors [30].

In this paper, we take advantage of county-level variation in Covid-19 mortality to estimate its relationship with all-cause mortality across US counties. We anticipate that counties with higher mortality from Covid-19 will also have experienced greater increases in mortality from other causes of death because the impact of the pandemic is not registered in Covid-19 deaths alone. We use the relationship between Covid-19 mortality and changing mortality from all causes of death to estimate excess mortality that was not directly assigned to Covid-19 as a cause of death. While prior studies generated a prediction of expected mortality in 2020 based on historical trends and then compared expected to observed mortality to estimate excess deaths, our model takes advantage of county variation to estimate simultaneously the mortality trend and the imprint of Covid-19. We then examine how the extent of excess mortality not assigned to Covid-19 varies across subsets of counties defined by area-level sociodemographic and health characteristics, allowing us to identify population subgroups with a disproportionate number of excess deaths that were not directly assigned to Covid-19. Our estimates provide an alternative approach to calculating excess deaths that can complement existing approaches.

## Methods

Our analytic plan, designed prior to reviewing our data or beginning analysis, was to estimate excess deaths in 2020 using the relationship between 2020 all-cause mortality, directly assigned Covid-19 mortality, and historical all-cause mortality at the county-level. Though we did not have a formal, prespecified plan, our a priori hypothesis was that excess deaths would exceed directly assigned Covid-19 deaths and that the percent of excess deaths not assigned to Covid-19 would vary by county-level factors. During the revision process, we introduced sensitivity analyses to assess the robustness of our results to alternative inclusion criteria, modified our approach to examining variation in excess deaths not assigned to Covid-19 to allow for full model stratification, and updated the analyses to reflect the most recent data and allow for a time lag that would better account for possible bias caused by incomplete data. The results of this study are reported as per the Strengthening the Reporting of Observational Studies in Epidemiology (STROBE) guidelines (S1 Table).

### Data Sources

We used NCHS provisional data on all-cause mortality and directly assigned Covid-19 mortality by county of residence from January 1 to December 31, 2020. The data were considered provisional due to a time lag between the occurrence of deaths and the completion, submission, and processing of death certificates. To account for possible lags in mortality reporting, we used data generated on March 12, 2021 (ten weeks after the end date), meaning that all deaths occurring before December 31 that were reported before March 12 were captured. Prior analyses of NCHS provisional data have found that provisional mortality data can have low completeness in the first month after a death occurs but are more than 75% complete within 8 weeks after a death [31]. 94% of deaths assigned to Covid-19 by NCHS had Covid-19 reported as the underlying cause of death; the other 6% had Covid-19 listed somewhere else on the death certificate [32]. The original NCHS data file included 3,140 counties, but the exact number of Covid-19 deaths was censored for counties with 1-9 deaths. We limited to counties with 20 or more Covid-19 deaths because death rate estimates based on death counts of less than 20 have a high relative standard error and are thus considered unreliable [33]. The final number of counties included in the analysis was 2,096, and the exclusion criteria are detailed in Fig S1.

To construct a historical comparison period, we utilized data from CDC Wonder reporting all-cause mortality for 2013 through 2018 by county of residence. We also used U.S. Census data on county population and age distribution from 2013 to 2020 and data on sociodemographic and health factors from a variety of consolidated sources including the 2020 RWJ Foundation County Health Rankings. A list of these data sources is provided in S2 Table. The present investigation relied on deidentified publicly available data and was therefore exempted from review by the Boston University Medical Center Institutional Review Board.

### Death Rates

We produced crude death rates for all-cause and directly coded Covid-19 mortality in 2020 using the reported death counts and the estimated county-level population on July 1, 2020. To compute an average historical death rate for 2013 to 2018, we divided the sum of deaths from 2013 to 2018 by the total population from 2013 to 2018.

### Sociodemographic and Health Factors

Prior literature has documented differences in Covid-19 mortality by sociodemographic and health factors [24,34]. In this analysis, the sociodemographic factors that we examined were age (% over 65 years), rurality (% rural), population distribution by race/ethnicity (% Hispanic, % non-Hispanic Black, % non-Hispanic White), socioeconomic status (median household income and % with some college or more education), and housing (% homeownership). For health factors, we examined poor health (% living with poor or fair health), obesity (% with obesity), smoking (% who smoke), and diabetes (% with diabetes). We stratified counties into population weighted quartiles for each sociodemographic or health factor, with particular attention to counties in the upper and lower 25% of values.

### Statistical Analysis

We first modeled all-cause mortality in 2020 as a function of historical all-cause mortality, directly assigned Covid-19 mortality in 2020, and an error term:

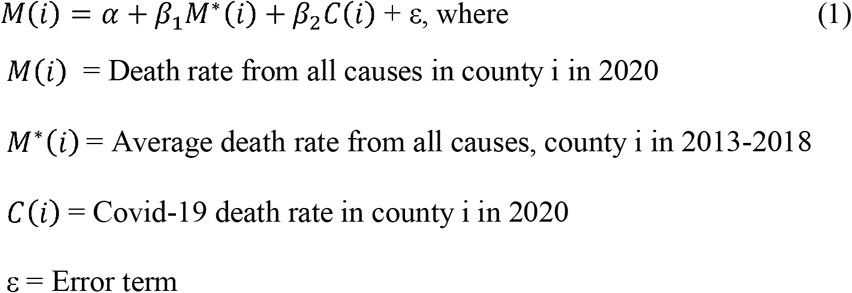

The parameters of equation (1) were estimated using Ordinary Least Squares regression with county units weighted by their population size. The value of *α* represents changes in mortality that are independent of Covid-19 mortality and that are common across regions. The value of *β*_1_ represents the extent to which past levels of all-cause mortality in a county are replicated in 2020. If *β*_1_ = 1.0, for example, then this would indicate that all-cause mortality in 2020 was on average equal to that in 2013-18, plus or minus the value of *α*. Together, combinations of *α* and *β*_1_ indicate how mortality changes that are not associated with Covid-19 vary with the level of all-cause mortality in 2013-18. The value of *β*_2_ indicates the extent to which mortality from Covid-19 affects all-cause mortality in 2020 after adjusting for historical mortality patterns. If *β*_2_ = 1.0, this would imply that each death coded to Covid-19 would be associated with one additional death from all causes combined. Values of *β*_2_ greater than 1.0 would suggest that the effect of the Covid-19 pandemic in a county is not fully reflected in deaths assigned to Covid-19 and that excess deaths are being attributed to some other causes of death. Values of *β*_2_ less than 1.0 would suggest that Covid-19 is over-recorded as a cause of death or reductions in mortality are occurring for other causes.

In secondary analyses, we limited our sample to counties with 50 or more Covid-19 deaths to assess the sensitivity of our results to counties reporting relatively small numbers of Covid-19 deaths. We also limited our sample to counties with populations greater than or equal to 50,000 to assess the robustness to excluding counties with relatively small populations. In further sensitivity analyses, we performed the regression among all counties in the original NCHS dataset, including counties that were eliminated by our study’s exclusion criteria. To assess the robustness of our results to alternative modeling approaches, we also estimated the relationship between Covid-19 and all-cause mortality using a Negative Binomial regression model.

We did not control for age in our primary analysis because the effect of age should be partially captured in the historical mortality term. In a sensitivity analysis, we re-estimated the primary OLS model using indirectly age-standardized death rates to adjust for differences in the counties’ age distributions. Deaths by age were not available at the county level so direct standardization of death rates was not possible. Indirect standardization adopts the age-specific death rate schedule for the whole US and applies it to the age distribution of a county to predict the number of deaths in that county [35]. It then calculates the ratio of actual deaths in the county to the predicted number of deaths. Finally, it applies that ratio to the US crude death rate to estimate the indirectly age-standardized death rate for the county [36]. Death rates and age distributions were employed in 10-year wide age intervals. When the death rate referred to Covid-19 mortality alone, death rates were restricted to that cause of death.

To identify county-level characteristics that modified the relationship between direct Covid-19 mortality and excess mortality (the *β*_2_ coefficient), we fully stratified our primary regression model into population weighted quartiles of various county-level sociodemographic and health factors. We then produced a forest plot displaying the *β*_2_ coefficients for the upper and lower 25% of each county-level factor. Counties with high *β*_2_ coefficients represent counties that have a high proportion of excess deaths not assigned to Covid-19 compared to their direct Covid-19 deaths. To understand how this relationship translated to absolute numbers of deaths, we also calculated predicted excess death rates in each of the strata using the fully stratified model coefficients. We determined the average observed directly assigned Covid-19 death rate for each stratum by calculating the weighted mean and then found the predicted excess death rate not assigned to Covid-19 by multiplying the average observed direct Covid-19 death rate for the stratum by the *β*_2_ coefficient minus 1. In a sensitivity analysis, we repeated both of these analyses using indirectly age standardized death rates.

## Results

Table 1 presents characteristics of the 2,096 counties included in the sample, whose distribution across the U.S. is visualized in S2 Fig. The total number of residents living in these counties was 319.1 million. Among these counties, 15.9% of the population was older than 65 years compared to 16.0% of the population in all counties in the U.S. The counties in the sample were 16.7% rural, which was lower than all counties in the U.S. which were 18.6% rural. On average, the counties in the sample were 18.7% Hispanic, 12.7% non-Hispanic Black and 59.6% non-Hispanic White. The median household income in the counties was $65,603. In the counties, 63.4% of residents were homeowners, and 16.7% were living with poor or fair health. The mean directly assigned Covid-19 death rate in 2020 for the sample was 1.2 deaths per 1000 person-years, and the mean all-cause death rate in 2020 was 10.1 deaths per 1000 person-years

**Table 1.**
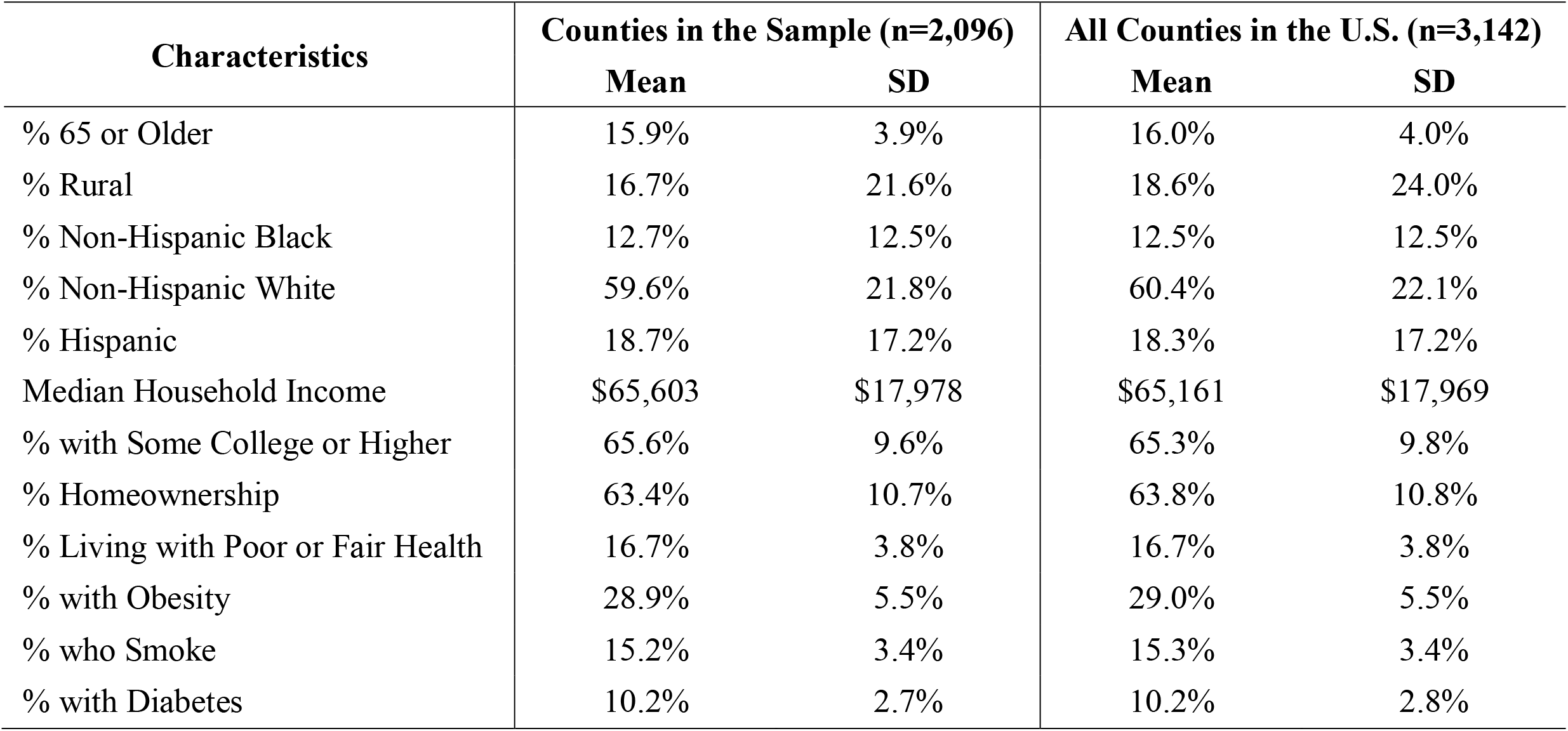
Characteristics of Counties in the Sample Compared to All Counties in the U.S.

Fig 1 plots the difference between the 2020 all-cause mortality rate and the average 2013-2018 historical all-cause death rate against the Covid-19 mortality rate in the study counties. The area of each point is roughly proportional to the county’s population size. This figure presents evidence that there is a positive relationship between the change in mortality from all causes of death and the level of Covid-19 mortality in a county. The slope of the regression line (solid blue) is steeper than the 45-degree line (dashed grey), indicating that one additional Covid-19 death is associated with more than one additional death from all causes.

**Fig 1.**
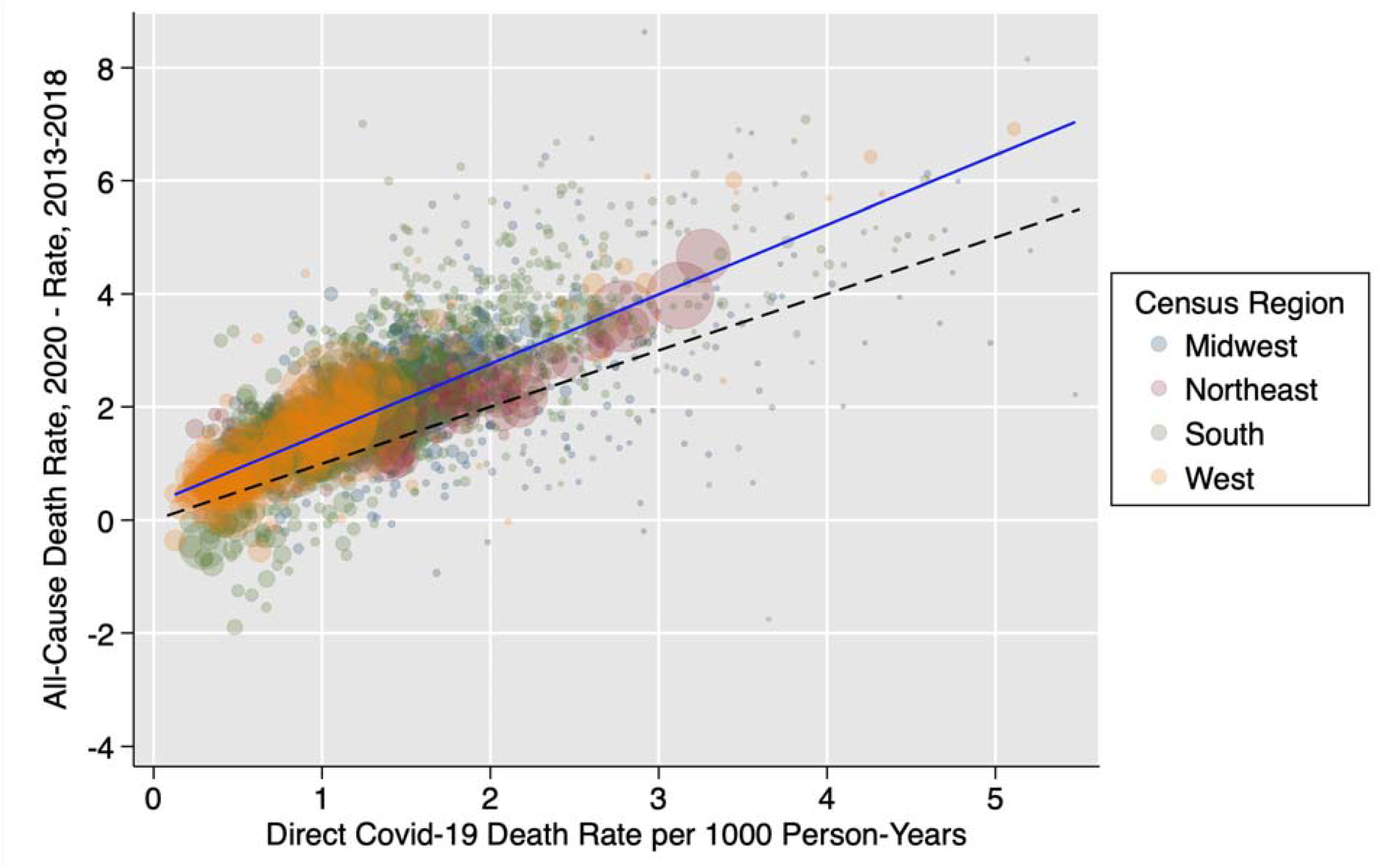
Difference Between the 2020 All-Cause Mortality Rate and 2013-2018 Historical Mortality Rate vs. 2020 Direct Covid-19 Mortality Rate (n=2,096)^a,b,c^ a. The solid blue line represents a linear model of best fit weighted by population size. b. The dashed reference line represents a slope of 1. c. Six counties with a direct Covid-19 death rate greater than 6 deaths per 1000 person-years or a difference between 2020 all-cause and 2013-2018 all-cause death rates greater than 10 deaths per 1000 person-years were excluded from the figure for the purposes of visualization. These counties were included in all regressions.

Table 2 presents coefficients of the model describing the relationship between the directly assigned Covid-19 mortality rate and all-cause mortality in 2020. The estimated value of *α* is 0.105 deaths per 1000 people and *β*_1_ is 1.03 (95% CI, 1.02 to 1.04). This combination suggests that crude all-cause mortality rose on average across counties between 2013-18 and 2020. The coefficient of *β*_2_ is estimated to be 1.20 (95% CI, 1.16 to 1.24). This value suggests that, for every 100 deaths assigned to Covid-19, the number of all-cause deaths rose by 120. This result implies that (120 – 100) / 120 or 17% (95% CI, 14% to 19%) of all excess deaths were not directly assigned to Covid-19 on death certificates. In absolute terms, 367,820 directly assigned Covid-19 deaths occurred in the 2,096 counties in the study between January 1 and December 31, 2020, meaning there were 74,172 (95% CI, 60,162 to 88,183) excess deaths not directly assigned to Covid-19 for a total of 441,992 (95% CI, 427,982 to 456,003) excess deaths.

**Table 2.**
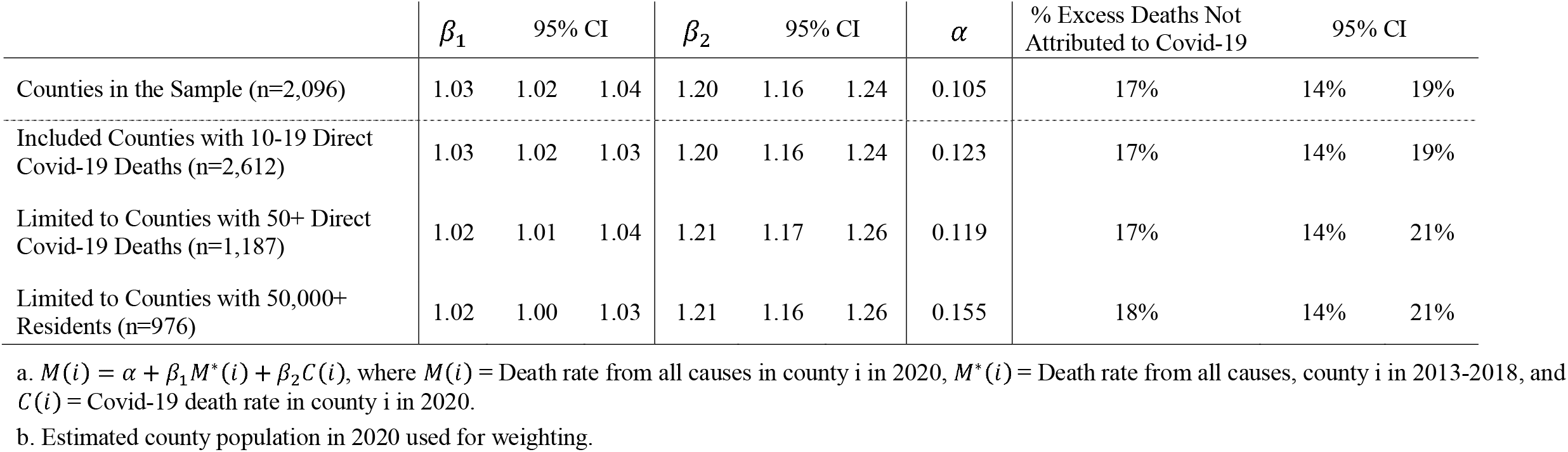
Estimated Parameters of Linear Models of the Relationship Between Directly Assigned Covid-19 Deaths, Historical All-Cause Mortality and All-Cause Mortality in 2020^a,b^

When we limited to counties with 50 or more direct Covid-19 deaths, the coefficient of *β*_2_ was 1.21 (95% CI, 1.17 to 1.26). *β*_2_ was 1.21 (95% CI, 1.16 to 1.26) when we limited to counties with 50,000 or more residents, and *β*_2_ was 1.20 (95% CI, 1.16 to 1.24) when we included counties with 10-19 Covid-19 deaths. Our results were relatively consistent with indirect age standardization, with the percent of excess deaths not attributed to Covid-19 equaling 13% (95% CI, 11% to 16%). When we used an alternative modeling approach (a Negative Binomial model), we found that 22% of excess deaths were not assigned to Covid-19, suggesting that our OLS results may be conservative (S3 Table).

Fig 2 examines how county-level characteristics modified the *β*_2_ coefficient (the relationship between the rate of direct Covid-19 deaths and the rate of excess deaths from all causes). The figure displays estimates of the *β*_2_ coefficient across models that were fully stratified into population weighted quartiles for various sociodemographic and health factors. For sociodemographic factors, the *β*_2_ coefficient was higher in counties with the lowest median household incomes (1.31 [95% CI, 1.25, 1.38]) and the least formal education (1.29 [95% CI, 1.23, 1.35]) than in counties with the highest median incomes (1.05 [95% CI, 0.94, 1.15]) and most education (1.03 [95% CI, 0.92, 1.14]). The *β*_2_ coefficient was also elevated in counties with more non-Hispanic Black residents (1.30 [95% CI, 1.23, 1.38]) and fewer non-Hispanic White residents (1.22 [95% CI, 1.16, 1.28]) compared to counties with fewer non-Hispanic Black residents (1.16 [95% CI, 1.10, 1.22]) and more non-Hispanic White residents (1.02 [95% CI, 0.94, 1.10]). Regarding health factors, the *β*_2_ coefficient was higher in counties with the most poor or fair health (1.23 [95% CI,1.17, 1.30]) and the most diabetes (1.31 [95% CI, 1.25, 1.38]) compared to counties with the least poor or fair health (0.97 [95% CI, 0.88, 1.06]) or the least diabetes (1.10 [95% CI, 1.02, 1.19]). Regionally, the *β*_2_ coefficient was elevated in the South (1.43 [95% CI, 1.35, 1.51]) and West (1.55 [95% CI, 1.45, 1.65]) compared to the Northeast (1.18 [95% CI, 1.08, 1.27]) and Midwest (0.89 [95% CI, 0.79, 0.99]). S3 Fig presents the stratified models calculated using indirectly age standardized death rates. S4 Table presents the boundaries for the upper and lower quartiles for each of the sociodemographic and health factors examined in these stratified analyses.

**Fig 2.**
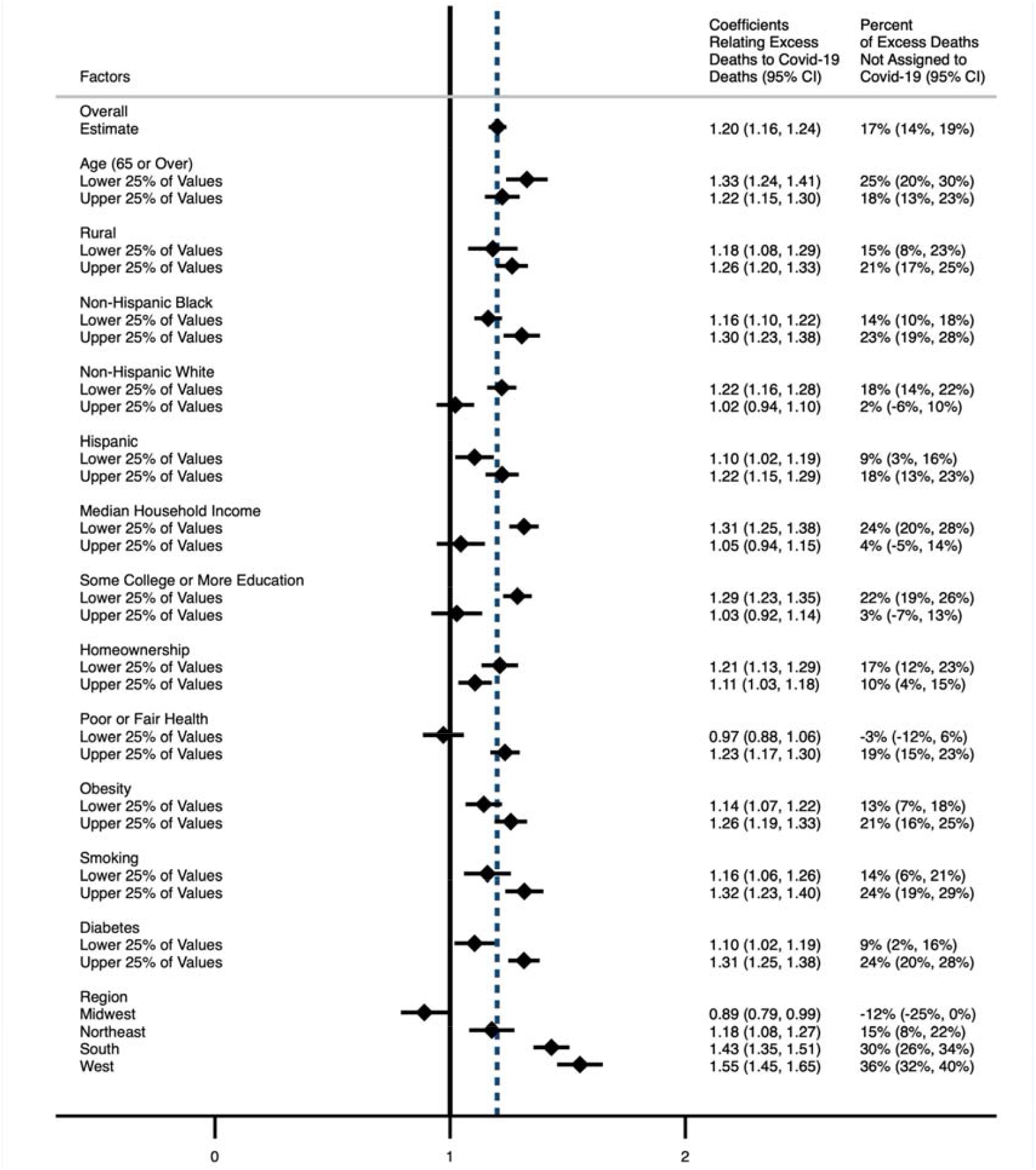
Relationship Between All-Cause Mortality and Direct Covid-19 Mortality across Strata of Sociodemographic and Health Factors^a,b,c,d,e^ a. n = 2,096 counties b. *β*_2_ coefficients generated from primary model:*M* (*i*) = *α β*_1_ *M*\* + *β*_2_*C*(*i*), where = Death rate from all causes in county i in 2020 *M\** (*i)*, = Death rate from all causes, county i in 2013-2018, and *C*(*i)* = Covid-19 death rate in county i in 2020. The model was weighted by the 2020 population and fully stratified into population weighted quartiles for each sociodemographic or health factor. The coefficients for the upper and lower 25% of values for each factor are presented in this figure. c. Sample interpretation: in counties with lower household income, for every 1 directly assigned Covid-19 death, there was an increase in 1.31 all-cause deaths, suggesting there were 0.31 deaths not assigned to Covid-19 for every 1 directly assigned Covid-19 death in these counties. d. Indirectly age-standardized models are presented in S3 Fig. e. The dashed line represents the overall estimate. The solid line represents a coefficient of 1, which indicates that for every 1 directly assigned Covid-19 deaths, 0 deaths not assigned to Covid-19 occurred.

Fig 3 decomposes the 2020 excess death rate into the observed excess death rate directly assigned to Covid-19 and the predicted excess death rate not assigned to Covid-19 and compares counties in the upper and lower 25% of county-level factors. Predictions were generated using the fully stratified models presented in Fig 2. S4 Fig presents decomposed death rates with indirect age standardization. These figures reveal that some counties reported high directly assigned Covid-19 mortality (i.e. counties in the Northeast and counties with low median household income), some had high excess mortality (i.e. counties in the South and West), and others experienced both high direct Covid-19 mortality and excess mortality (i.e. counties with the most non-Hispanic Black residents). Comparing counties with the most non-Hispanic Black residents to counties with the fewest non-Hispanic Black residents, substantial racial inequities were observed in direct Covid-19 mortality and in excess deaths not assigned to Covid-19.

**Fig 3.**
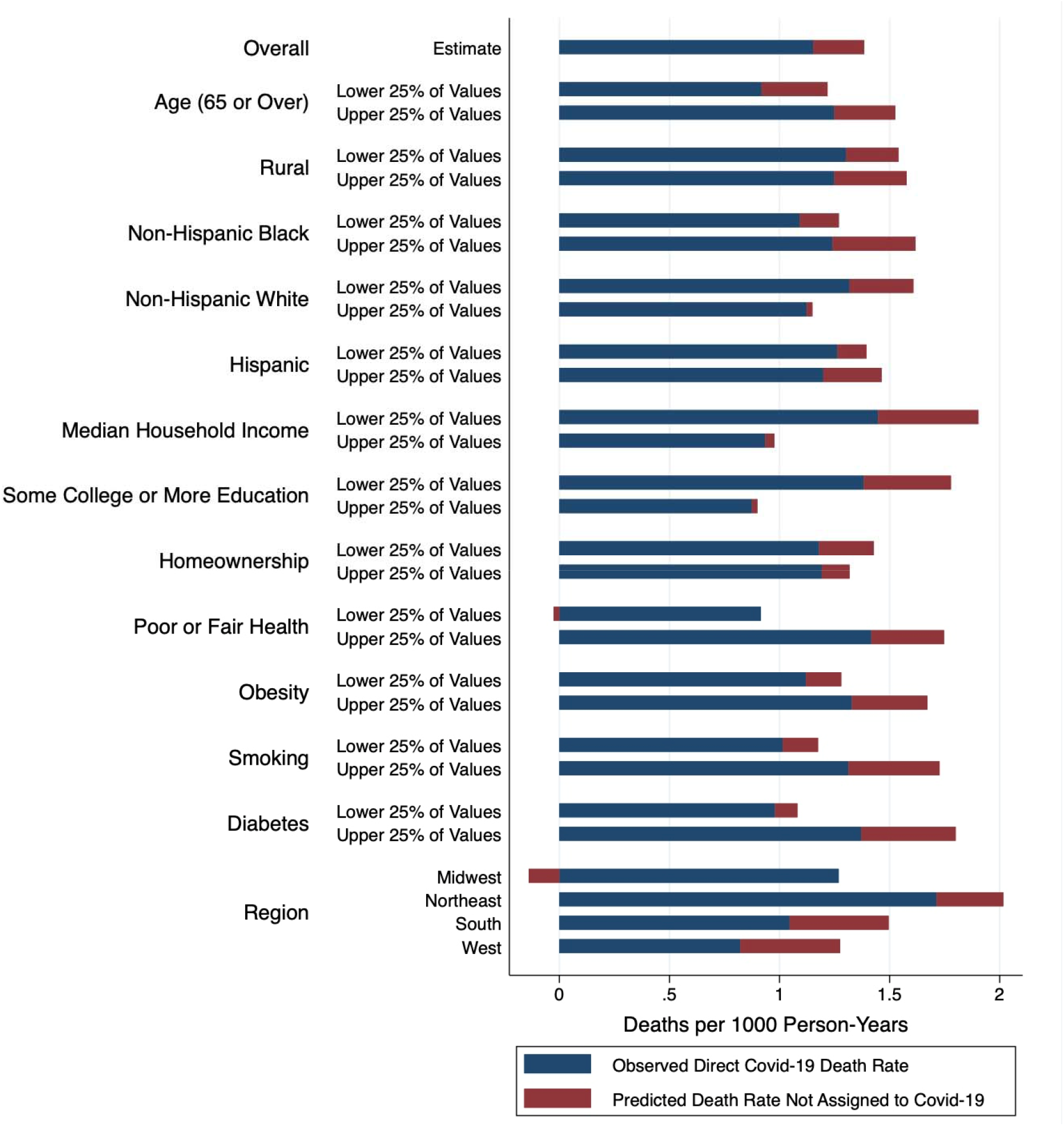
Decomposition of 2020 Excess Death Rates across Strata of Sociodemographic and Health Factors^a,b,c^ a. n = 2,096 counties b. Predicted death rates generated from primary model: *M* (*i*) = *α β*_1_ *M*\* (*i*) + *β*_2_*C*(*i*), where *M* (*i)* = Death rate from all causes in county i in 2020, *M\** (*i*) = Death rate from all causes, county i in 2013-2018, and *C*(*i*) = Covid-19 death rate in county i in 2020. The model was weighted by the 2020 population and fully stratified into population weighted quartiles for each sociodemographic or health factor. The death rates for the upper and lower 25% of values for each factor are presented in this figure. c. Indirectly age-standardized death rates are presented in S4 Fig.

## Discussion

In this study, we estimated that 17% of excess deaths attributable to the Covid-19 pandemic were not assigned to Covid-19 on death certificates. The proportion of excess deaths not counted as direct Covid-19 deaths was even higher in counties with lower average socioeconomic status, counties with more comorbidities, and counties in the South and West. Counties with more non-Hispanic Black residents, who were already at high risk of Covid-19 death based on direct counts, also reported a higher percentage of excess deaths not assigned to Covid-19, indicating a pattern related to structural racism.

Prior estimates of excess mortality not assigned to Covid-19 have been relatively similar despite being based on different methodological approaches. Weinberger et al. identified 95,235 directly coded Covid-19 deaths from March 1 to May 30, 2020, and 122,300 total excess deaths attributable to the pandemic [21]. This indicated that 27,065 deaths or 22% of excess deaths were excess deaths not assigned to Covid-19. Additionally, the NCHS has identified 288,287 directly coded Covid-19 deaths and between 291,208 and 400,791 total excess deaths as of December 22, 2020 [39]. This data suggests that between 1% and 28% of excess deaths were not directly assigned to Covid-19. An analysis by Woolf et al., based on data from March 1 through April 25, 2020, yielded a higher estimate, finding that of 87,001 excess deaths, 30,755 excess deaths or 35% were not assigned to Covid-19 [22]. A subsequent analysis by Woolf et al. using data from March 1 through August 1 found that of 225,530 excess deaths, 150,541 were directly assigned to Covid-19, suggesting that 33% of excess deaths were not assigned to Covid-19 [37].

The coefficients from our primary model indicate that mortality would have risen between 2013-18 and 2020 even in the absence of the Covid-19 pandemic. This prediction is consistent with a rising trend in national crude death rates between 2013 and 2018. The crude death rate rose from 821.5 deaths per 100,000 people in the year 2013 to 867.8 deaths per 100,000 people in 2018 [40]. Estimates of excess deaths that fail to account for these increases, which are likely attributable primarily to population aging, may result in overestimates of the percentage of excess deaths not assigned to Covid-19 [41].

As noted earlier, excess deaths not assigned to Covid-19 could include deaths involving Covid-19 that were misclassified to other causes of death and deaths indirectly related to the Covid-19 pandemic. The NCHS has examined excess deaths not assigned to Covid-19 by cause of death nationally. As of December 22, 2020, NCHS has attributed 38,115 excess deaths to Alzheimer’s disease and related dementias, 22,661 excess deaths to hypertensive diseases, 14,684 excess deaths to ischemic heart disease, 14,194 excess deaths to diabetes, and 3,060 excess deaths to influenza and pneumonia [39]. It is possible that a substantial fraction of the deaths of individuals with pre-existing chronic conditions who acquire Covid-19 and die as a result are ascribed to the pre-existing condition. These may constitute many of the excess deaths not attributed to Covid-19. This explanation is consistent with our finding that excess mortality not assigned to Covid-19 was higher in counties with higher levels of poor health and diabetes.

In this study, we found that counties with low median incomes and less formal education, and counties in the South and West reported high numbers of excess deaths not assigned to Covid-19 compared to direct Covid-19 deaths, suggesting that Covid-19 mortality may be especially undercounted in these areas. Determining the cause of potential undercounting is an important area for future research. Possible factors could include lower rates of Covid-19 testing in these populations [42], reduced access to health care [43,44], regional diagnostic and coding differences [45], or political attitudes about the Covid-19 pandemic [46,47].

Indirect deaths, such as suicide, homicide, or drug overdose and deaths related to reductions in health care use, may also be higher in these areas. Faust et al. found that only 38% of excess deaths in 2020 among adults aged 25 to 44 years were attributed to Covid-19, suggesting substantial mortality among younger adults during the pandemic that is not accounted for in Covid-19 death tallies [48]. Analyses of multiple cause-of-death data, when available, will help to shed additional light on the contribution of Covid-19 to US mortality and help to explain why excess all-cause mortality in the U.S. is high compared to other countries including those with high directly assigned Covid-19 mortality [49].

Previous research has shown that counties with a higher percentage of Black residents have reported more mortality attributable to Covid-19 [24,50]. According to underlying cause of death data, Black people are 1.7 times more likely to die from Covid-19 than White people [51]. Racial inequities in Covid-19 mortality relate to structural racism that has made Black people more likely to be exposed to Covid-19 at work, in transportation, and in housing during the pandemic [24,36,50,52]. Another factor is racial health inequities in asthma, COPD, hypertension and diabetes, which are risk factors for Covid-19 [53,54]. The presence of these comorbidities could also reduce the likelihood of assigning Covid-19 as a cause of death. Our analysis suggests that the substantial racial inequities observed in directly assigned Covid-19 death rates for the non-Hispanic Black population are even larger in excess death rates not assigned to Covid-19.

Our analysis had several limitations. The 2020 all-cause mortality and Covid-19 mortality data used were provisional. Counties may have differential delays in reporting death certificate data that vary by county, state, rurality, or other area-level factors. In particular, counties that are currently reporting lower all-cause mortality in 2020 than in the historical period are notable. While it is possible that these counties represent random annual variation in death rates or incomplete data, these counties could also have experienced reductions in mortality as a result of strong public health measures that reduced other causes of death such as influenza [19]. Future research should investigate this subset of counties in more detail to understand what led to their apparent success relative to other counties. If differential reporting delays occurred for direct Covid-19 mortality but not for all-cause mortality, we may have overestimated the percent of excess deaths that would not be assigned to Covid-19 when final data are available. To address this potential limitation, we used data that had a ten week lag, meaning that all deaths occurring before December 31, 2020 which were reported and processed before March 12, 2021 were included. Another potential source of error in our analysis is that death rates calculated from small numbers of cases may be less accurate than death rates calculated for areas with larger numbers of cases. To account for this possibility, we limited our dataset to counties with 20 or more direct Covid-19 deaths. We also conducted sensitivity analyses where we limited to counties with 50,000 or more residents and restricted to counties with 50 or more direct Covid-19 deaths. Results were not sensitive to these alternative restrictions. Our analytic approach may also be affected by measurement error in Covid-19 mortality in a way that differs from its effect on other estimates. In general, random measurement error in an independent variable (such as Covid-19 mortality in our analysis) is expected to bias the coefficient of that variable towards the null, or zero [55]. If our estimate of *β*_2_ is biased downwards, then we will have underestimated the magnitude of excess deaths. Another limitation of this study is that we lacked disaggregated data on county-level mortality by age, sex, race/ethnicity, and sociodemographic and health characteristics. As a result, age-compositional effects were addressed using an indirect age standardization procedure and predictors were assessed based on their distributions at the county-level. Future research based on county-level data with further disaggregation is needed to confirm our findings. Lastly, the sociodemographic and health factors examined in this analysis were based on data from 2010 through 2018. County-level distribution of these factors may have changed between that time and 2020 when the mortality data was analyzed.

## Conclusion

In line with previously published studies, our findings suggest that the overall mortality burden of Covid-19 considerably exceeds reported Covid-19 deaths. Using provisional vital statistics county-level data on Covid-19 and all-cause mortality in 2020 from the NCHS, we estimated that 17% of all excess deaths in the United States from January 1 to December 31, 2020 were excess deaths not assigned to Covid-19. Racial and socioeconomic inequities in Covid-19 mortality also increased when excess deaths not assigned to Covid-19 were considered. Our findings emphasize the importance of considering health equity in the policy response to the pandemic, such as in access to Covid-19 vaccines. As the impact of Covid-19 continues to spread in the U.S., analysis of excess deaths will continue to be a valuable proxy measure for assessing the overall mortality burden of Covid-19. This study highlights the importance of considering excess deaths beyond those directly assigned to Covid-19 in the overall assessment of the mortality impact of the Covid-19 pandemic and provides a new method for doing so.

## Data Availability

All data used in this manuscript are publicly available with the exception of the 2020 county population data, which are available through a special request to the U.S. Census Bureau. Further details about the data used in this analysis are provided at the linked GitHub repository.

https://github.com/pophealthdeterminantslab/covid-19-county-analysis

## Acknowledgements

The Robert Wood Johnson Foundation supported the research reported in this publication (Grant #77521). ITE was also supported by National Institute on Aging R01 AG060115 “Causes of Geographic Divergence in American Mortality Between 1990 and 2015: Health Behaviors, Health Care Access and Migration.”

The authors would like to thank Farida Ahmad, Robert Anderson, Magali Barbieri, Courtney Boen, Dana Glei, Josh Goldstein, Michelle Guillot, Patrick Heuveline, Anna McGregor, Jennifer Weuve and Wubin Xie for their valuable feedback on the manuscript.

## Disclaimer

The interpretations, conclusions, and recommendations in this work are those of the authors and do not necessarily represent the views of the Robert Wood Johnson Foundation.

