## Supplementary Materials for "Assessing the Impact of the Covid-19 Pandemic on US Mortality: A County-Level Analysis"

**S1 Table.** STROBE Checklist

| | Item No | Recommendation | Section, Paragraph |
|---|---|---|---|
| **Title and abstract** | 1 | (*a*) Indicate the study's design with a commonly used term in the title or the abstract | Abstract, Paragraph 2 |
| | | (*b*) Provide in the abstract an informative and balanced summary of what was done and what was found | Abstract, Paragraph 2 |
| **Introduction** | | | |
| Background/rationale | 2 | Explain the scientific background and rationale for the investigation being reported | Introduction, Paragraphs 2-4 |
| Objectives | 3 | State specific objectives, including any prespecified hypotheses | Introduction, Paragraph 5 |
| **Methods** | | | |
| Study design | 4 | Present key elements of study design early in the paper | Methods, Paragraph 2-3 |
| Setting | 5 | Describe the setting, locations, and relevant dates, including periods of recruitment, exposure, follow-up, and data collection | Methods, Paragraph 2-3 |
| Participants | 6 | (*a*) *Cohort study*—Give the eligibility criteria, and the sources and methods of selection of participants. Describe methods of follow-up<br>*Case-control study*—Give the eligibility criteria, and the sources and methods of case ascertainment and control selection. Give the rationale for the choice of cases and controls<br>*Cross-sectional study*—Give the eligibility criteria, and the sources and methods of selection of participants | Methods, Paragraph 2 |
| | | (*b*) *Cohort study*—For matched studies, give matching criteria and number of exposed and unexposed<br>*Case-control study*—For matched studies, give matching criteria and the number of controls per case | n/a |
| Variables | 7 | Clearly define all outcomes, exposures, predictors, potential confounders, and effect modifiers. Give diagnostic criteria, if applicable | Methods, Paragraphs 3-5 |
| Data sources/ measurement | 8* | For each variable of interest, give sources of data and details of methods of assessment (measurement). Describe comparability of assessment methods if there is more than one group | Methods, Paragraphs 3-5 |
| Bias | 9 | Describe any efforts to address potential sources of bias | Methods, Paragraphs 9-10 |
| Study size | 10 | Explain how the study size was arrived at | Methods, Paragraph 2 |
| Quantitative variables | 11 | Explain how quantitative variables were handled in the analyses. If applicable, describe which groupings were chosen and why | Methods, Paragraphs 4-5 |
| Statistical methods | 12 | (*a*) Describe all statistical methods, including those used to control for confounding | Methods, Paragraphs 6-7 |
| | | (*b*) Describe any methods used to examine subgroups and interactions | Methods, Paragraph 10 |
| | | (*c*) Explain how missing data were addressed | Methods, Paragraph 8 |
| | | (*d*) *Cohort study*—If applicable, explain how loss to follow-up was addressed | n/a |

| | | | |
|---|---|---|---|
| | | *Case-control study*—If applicable, explain how matching of cases and controls was addressed<br>*Cross-sectional study*—If applicable, describe analytical methods taking account of sampling strategy | |
| | | (*e*) Describe any sensitivity analyses | Methods, Paragraphs 8-9 |

**Results**

| | | | |
|---|---|---|---|
| Participants | 13* | (a) Report numbers of individuals at each stage of study—eg numbers potentially eligible, examined for eligibility, confirmed eligible, included in the study, completing follow-up, and analysed | Methods, Paragraph 2 Results, Paragraph 1 |
| | | (b) Give reasons for non-participation at each stage | Methods, Paragraph 2 |
| | | (c) Consider use of a flow diagram | S1 Fig |
| Descriptive data | 14* | (a) Give characteristics of study participants (eg demographic, clinical, social) and information on exposures and potential confounders | Table 1 |
| | | (b) Indicate number of participants with missing data for each variable of interest | S1 Fig |
| | | (c) *Cohort study*—Summarise follow-up time (eg, average and total amount) | n/a |
| Outcome data | 15* | *Cohort study*—Report numbers of outcome events or summary measures over time | n/a |
| | | *Case-control study*—Report numbers in each exposure category, or summary measures of exposure | n/a |
| | | *Cross-sectional study*—Report numbers of outcome events or summary measures | Results, Paragraph 1 |
| Main results | 16 | (*a*) Give unadjusted estimates and, if applicable, confounder-adjusted estimates and their precision (eg, 95% confidence interval). Make clear which confounders were adjusted for and why they were included | Results, Paragraph 3-4 |
| | | (*b*) Report category boundaries when continuous variables were categorized | S4 Table |
| | | (*c*) If relevant, consider translating estimates of relative risk into absolute risk for a meaningful time period | Results, Paragraph 3 |
| Other analyses | 17 | Report other analyses done—eg analyses of subgroups and interactions, and sensitivity analyses | Results, Paragraphs 4-6 |

**Discussion**

| | | | |
|---|---|---|---|
| Key results | 18 | Summarise key results with reference to study objectives | Discussion, Paragraphs 1, 5, 7 |
| Limitations | 19 | Discuss limitations of the study, taking into account sources of potential bias or imprecision. Discuss both direction and magnitude of any potential bias | Discussion, Paragraph 8 |
| Interpretation | 20 | Give a cautious overall interpretation of results considering objectives, limitations, multiplicity of analyses, results from similar studies, and other relevant evidence | Discussion, Paragraphs 1-7 |
| Generalisability | 21 | Discuss the generalisability (external validity) of the study results | Discussion, Paragraph 8 |

**Other information**

| | | | |
|---|---|---|---|
| Funding | 22 | Give the source of funding and the role of the funders for the present study and, if applicable, for the original study on which the present article is based | Acknowledgments |

**S2 Table**. Data Sources and Years for County-Level Factors

| **Variable** | **Data Source:** |
|---|---|
| % 65 Years and Older | Census Population Estimates, 2018 |
| % Rural | Census Population Estimates, 2010 |
| % Hispanic | Census Population Estimates, 2018 |
| % Non-Hispanic Black | Census Population Estimates, 2018 |
| % Non-Hispanic White | Census Population Estimates, 2018 |
| Median Household Income | Small Area Income and Poverty Estimates, 2018 |
| % with Some College or Higher | American Community Survey, 5-year estimates, 2014-2018 |
| % Homeownership | American Community Survey, 5-year estimates, 2014-2018 |
| % with Poor or Fair Health | Behavioral Risk Factor Surveillance System, 2017 |
| % with Obesity | United States Diabetes Surveillance System, 2016 |
| % who Smoke | Behavioral Risk Factor Surveillance System, 2017 |
| % with Diabetes | United States Diabetes Surveillance System, 2016 |

**S3 Table**. Comparison of OLS, Indirectly Age Standardized and Negative Binomial Models

| Model | Number of Excess Deaths per 1 Directly Coded Covid-19 Death | % Excess Deaths Not Attributed to Covid-19 |
|---|---|---|
| OLS Model[a,b] | 1.20 [95% CI (1.16, 1.24)] | 17% [95% CI (14%, 19%)] |
| OLS Model, Age-Standardized[a,b,c] | 1.15 [95% CI (1.12, 1.19)] | 13% [95% CI (11%, 16%)] |
| Negative Binomial Model[d,e] | 1.28 | 22% |

a. The OLS models were specified as $M(i) = \alpha + \beta_1 M^*(i) + \beta_2 C(i)$, where $M(i)$ = Death rate from all causes in county i in 2020, $M^*(i)$ = Death rate from all causes, county i in 2013-2018, and $C(i)$ = Covid-19 death rate in county i in 2020. Model weighted by the 2020 population. For the Negative Binomial model, $M(i) =$ deaths from all-causes in county i in 2020 rather than the death rate, with the 2020 population used as an offset.

b. Number of excess deaths per 1 directly coded Covid-19 death is equivalent to the regression coefficient for directly coded Covid-19 deaths.

c. Death rates were indirectly age-standardized.

d. To calculate the number of excess deaths per 1 directly coded Covid-19 deaths, we used marginal prediction to calculate the all-cause death rate in 2020 at values of directly coded Covid-19 mortality that were +/- 0.1 deaths per 1000 people from the weighted mean of directly coded Covid-19 mortality. The change in all-cause mortality between these values was divided by 0.2 deaths per 1000 people to yield the number of excess deaths per 1 directly coded Covid-19 death.

e. A Poisson model was tested prior to the Negative Binomial model but was rejected due to poor goodness of fit.

**S4 Table.** Boundaries for Sociodemographic and Health Characteristic Quartiles[a]

| Characteristics | Lower 25% Quartile | | Upper 25% Quartile | |
|---|---|---|---|---|
| | Lowest Value | Highest Value | Lowest Value | Highest Value |
| % 65 or Older | 7.4% | 13.6% | 17.6% | 57.6% |
| % Rural | 0% | 1.3% | 24.3% | 100% |
| % Hispanic | 0.6% | 5.7% | 26.0% | 96.4% |
| % Non-Hispanic Black | 0.1% | 3.6% | 18.6% | 85.4% |
| % Non-Hispanic White | 2.7% | 42.1% | 77.9% | 97.9% |
| Median Household Income | 25,385 | 52,577 | 74,686 | 140,382 |
| % with Some College or Higher | 20.4% | 60.3% | 71.8% | 90.3% |
| % Homeownership | 19.6% | 56.9% | 71.0% | 89.8% |
| % Living with Poor or Fair Health | 8.1% | 14.0% | 18.9% | 41.0% |
| % with Obesity | 14.4% | 24.9% | 32.8% | 51.0% |
| % who Smoke | 5.9% | 12.6% | 17.6% | 41.5% |
| % with Diabetes | 2.9% | 8.4% | 11.5% | 34.1% |

a. Quartiles are weighted by the estimated 2020 population.

**S1 Fig**. Flowchart Detailing Sample Exclusions

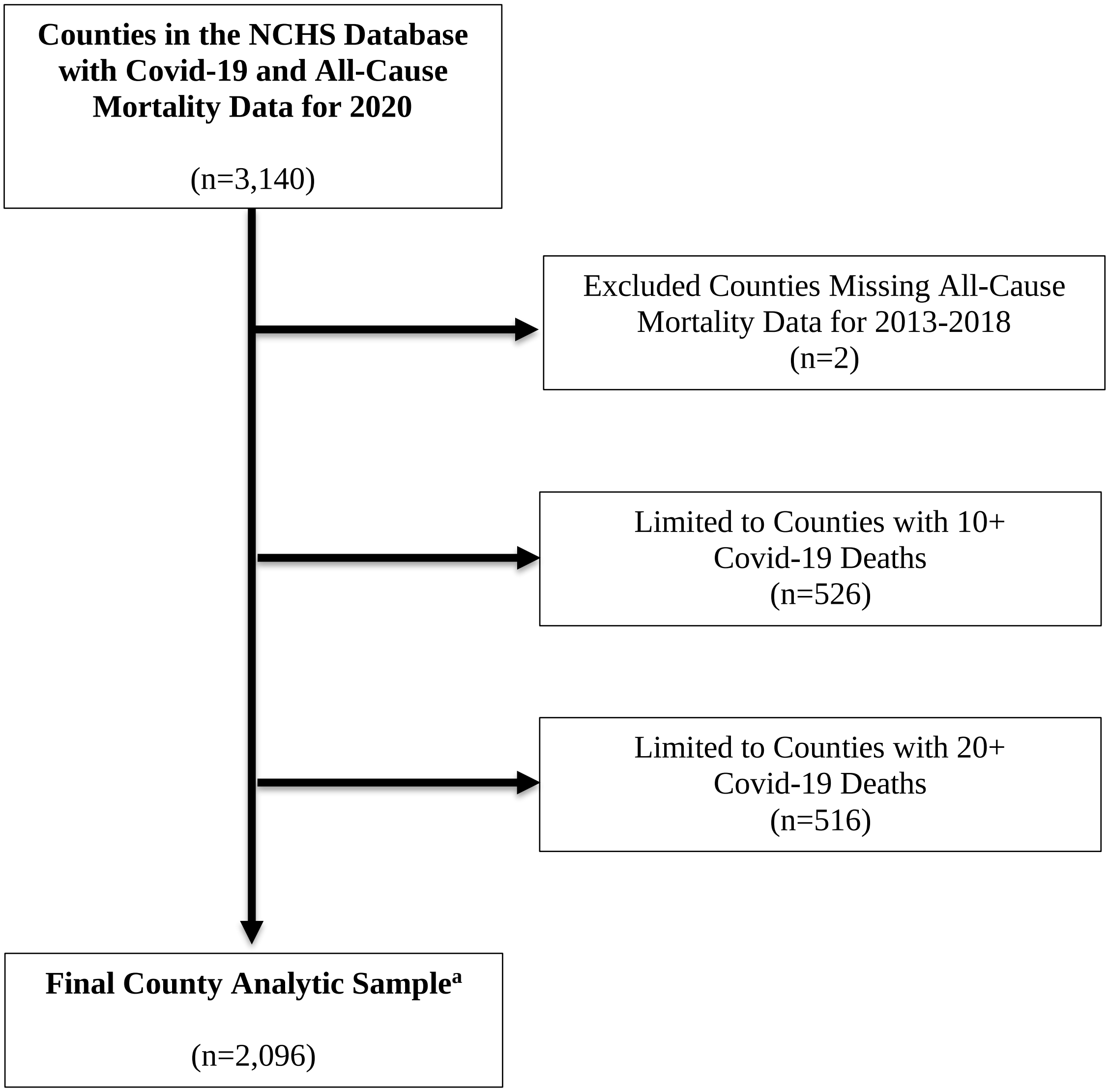


a. Counties had complete data for each of the sociodemographic and health characteristics we examined.

**S2 Fig**. US County Map Showing Geographic Distribution of Sample Counties (n=2,096)[a]

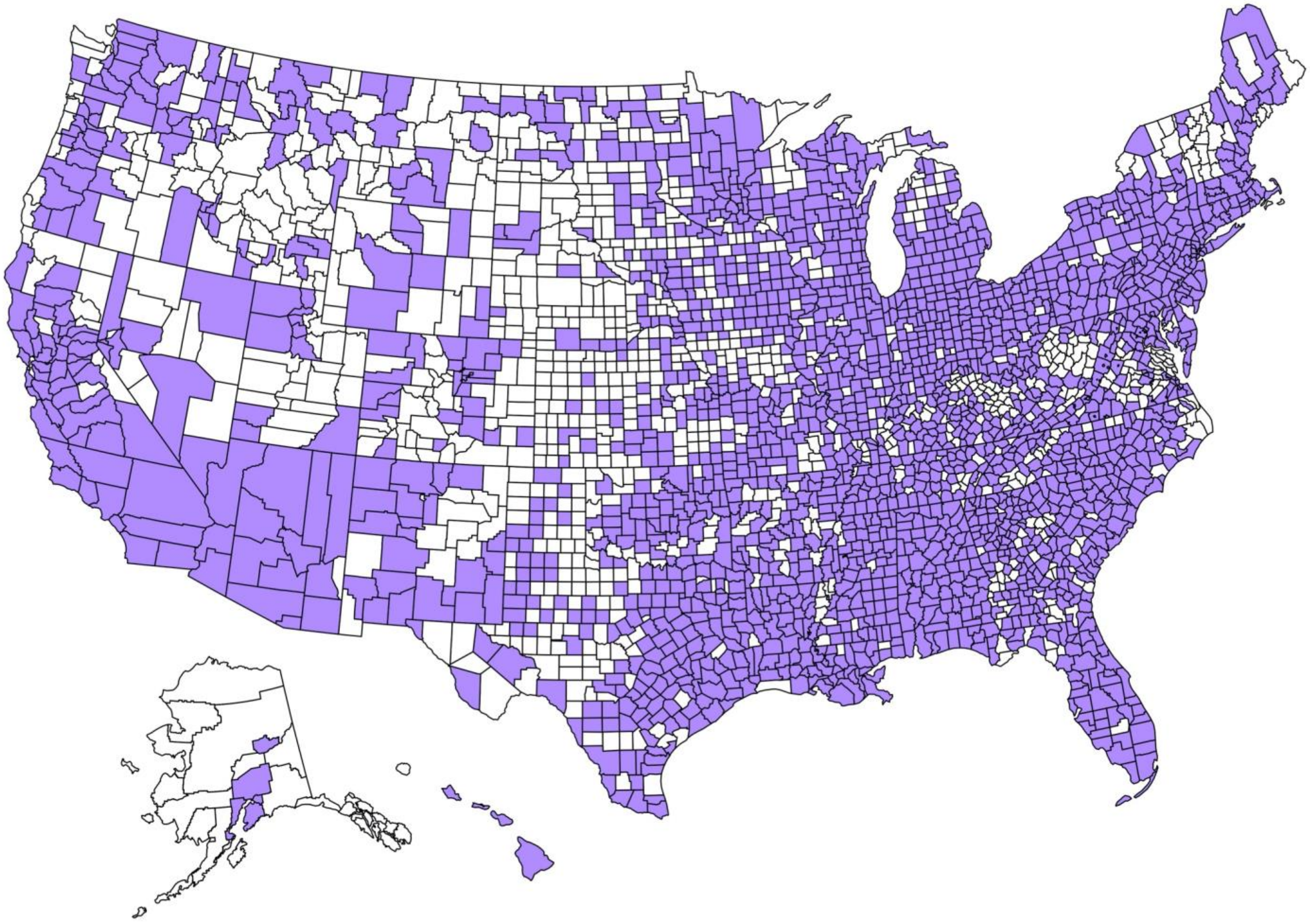

a. Map created using the usmap package in RStudio (https://CRAN.R-project.org/package=usmap)

**S3 Fig.** Relationship Between Indirectly Age Standardized All-Cause Mortality and Direct Covid-19 Mortality across Strata of Sociodemographic and Health Factors[a,b,c]

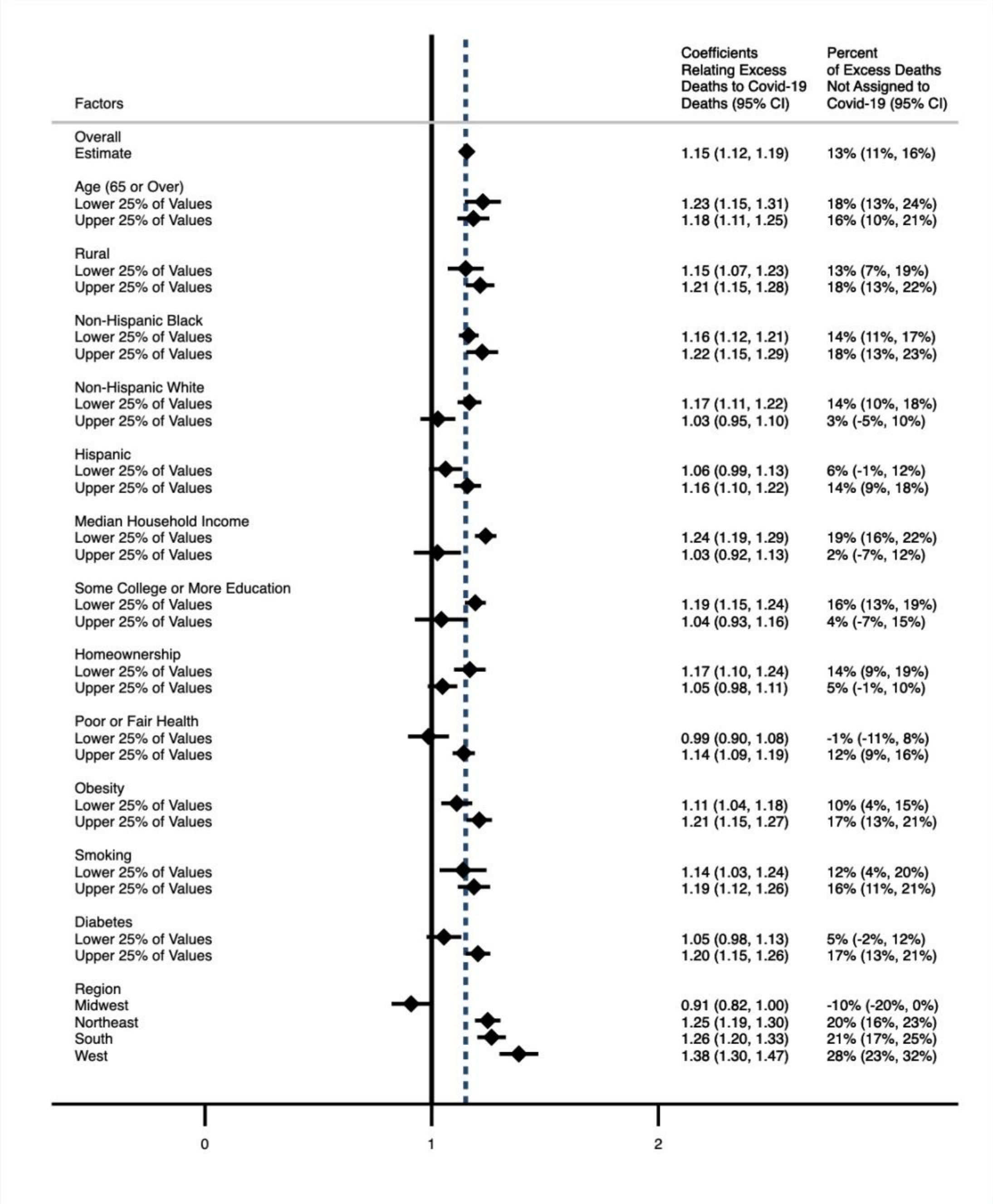


a. n = 2,096 counties
b. $\beta_2$ coefficients generated from primary model: $M(i) = \alpha + \beta_1 M^*(i) + \beta_2 C(i)$, where $M(i)$ = Death rate from all causes in county i in 2020, $M^*(i)$ = Death rate from all causes, county i in 2013-2018, and $C(i)$ = Covid-19 death rate in county i in 2020. The model was weighted by the 2020 population and fully stratified into population weighted quartiles for each sociodemographic or health factor. The coefficients for the upper and lower 25% of values for each factor are presented in this figure.
c. Sample interpretation: in counties with lower household income, for every 1 directly assigned Covid-19 death, there was an increase in 1.24 all-cause deaths, suggesting there were 0.24 deaths not assigned to Covid-19 for every 1 directly assigned Covid-19 death in these counties.

**S4 Fig.** Decomposition of 2020 Indirectly Age Standardized Excess Death Rates across Strata of Sociodemographic and Health Factors[a,b]

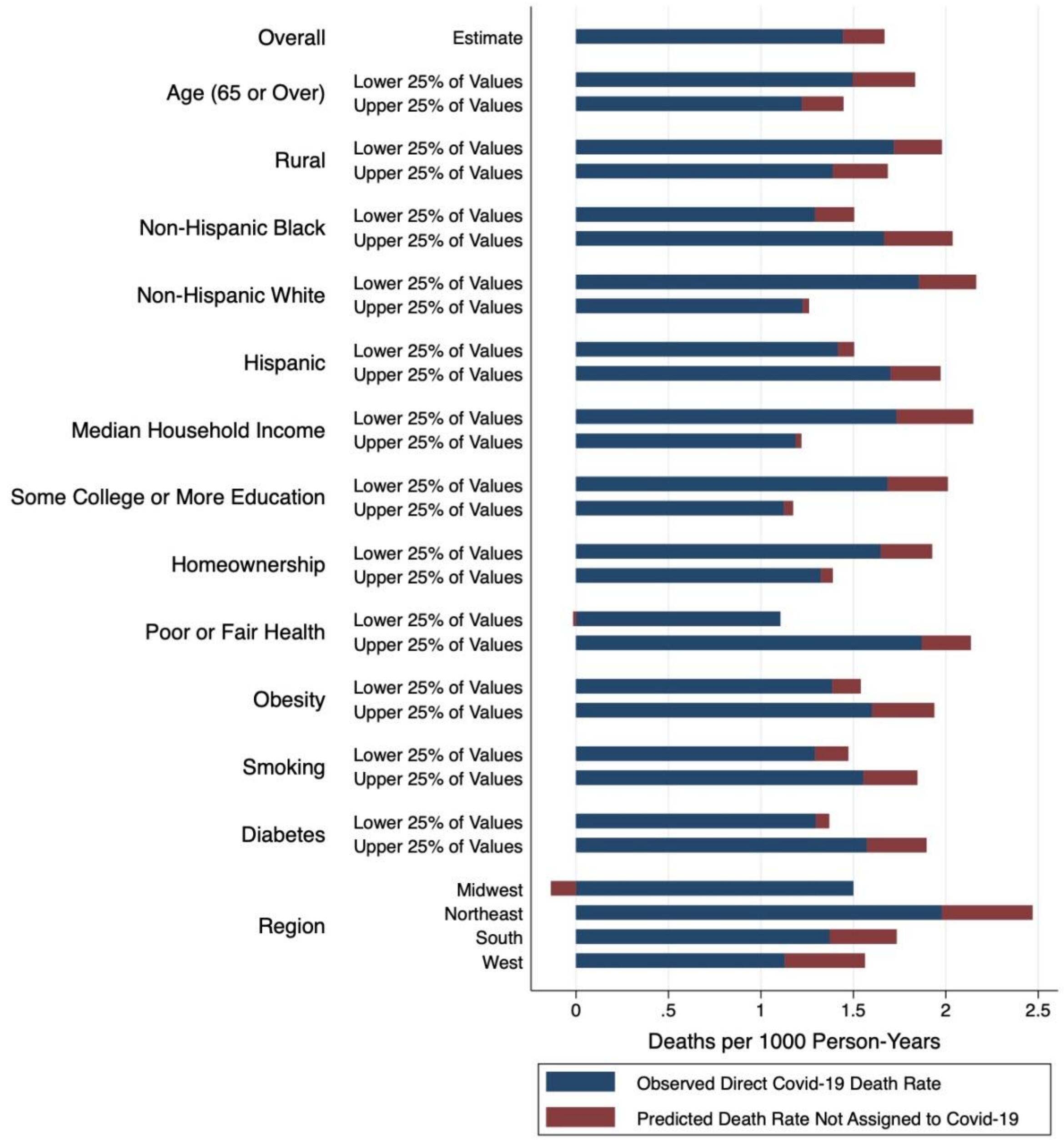


a. n = 2,096 counties

b. Predicted death rates generated from primary model: $M(i) = \alpha + \beta_1 M^*(i) + \beta_2 C(i)$, where $M(i)$ = Death rate from all causes in county i in 2020, $M^*(i)$ = Death rate from all causes, county i in 2013-2018, and $C(i)$ = Covid-19 death rate in county i in 2020. The model was weighted by the 2020 population and fully stratified into population weighted quartiles for each sociodemographic or health factor. The death rates for the upper and lower 25% of values for each factor are presented in this figure.
